# Bridging the Gap in Health Literacy: Harnessing the Power of Large Language Models to Generate Plain Language Summaries from Biomedical Texts

**DOI:** 10.1101/2024.07.02.24309847

**Authors:** Carolina Salazar-Lara, Andrés Felipe Arias Russi, Rubén Manrique

**Author notes:** Corresponding author: (CS).

## Abstract

Health literacy is essential for individuals to navigate the healthcare system and make informed decisions about their health. Low health literacy levels have been associated with negative health outcomes, particularly among older populations and those financially restricted or with lower educational attainment. Plain language summaries (PLS) are an effective tool to bridge the gap in health literacy by simplifying content found in biomedical and clinical documents, in turn, allowing the general audience to truly understand health-related documentation. However, translating biomedical texts to PLS is time-consuming and challenging, for which they are rarely accessible by those who need them. We assessed the performance of Natural Language Processing (NLP) for systematizing plain language identification and Large Language Models (LLMs), Generative Pre-trained Transformer (GPT) 3.5 and GPT 4, for automating PLS generation from biomedical texts. The classification model achieved high precision (97·2%) in identifying if a text is written in plain language. GPT 4, a state-of-the-art LLM, successfully generated PLS that were semantically equivalent to those generated by domain experts and which were rated high in accuracy, readability, completeness, and usefulness. Our findings demonstrate the value of using LLMs and NLP to translate biomedical texts into plain language summaries, and their potential to be used as a supporting tool for healthcare stakeholders to empower patients and the general audience to understand healthcare information and make informed healthcare decisions.

## Introduction

Health literacy refers to an individual’s capacity to access, understand, and use health information [1]. It empowers patients and their families to navigate healthcare systems, comprehend and act upon a diagnosis or medical instruction, adhere to medication regimens, and make informed decisions, otherwise considered daunting, regarding participation in clinical trials, treatment options, or medical procedures [2–4]. Low health literacy levels have been consistently associated with higher mortality rates, increased instances of preventable hospitalizations, and poor treatment adherence [3]. Paradoxically, while health literacy is crucial for positive health outcomes, the 2015 European Health Literacy Survey revealed that almost half of the respondents had inadequate health literacy, particularly among older populations, those who are financially restricted, or who have lower educational attainment [5–6].

With the growing expectation for individuals to participate in healthcare decisions, enhancing health literacy becomes a significant attribute in improving public health and reducing health disparities [1, 7–8].

Improving health literacy in the population extend beyond actions taken to increase individual health literacy levels. In line with the General Data Protection Regulation (GDPR) principle of transparency, stakeholders such as healthcare providers, policymakers, and pharmaceutical companies should strategize to improve their organizational health literacy (OHL) by ensuring the clarity and comprehensibility of health documentation [9–10].

One strategy to do so is by simplifying clinical and scientific research language into lay-friendly summaries, known as plain language summaries (PLS).

There are different techniques and guidelines that can be used to translate complex scientific and biomedical concepts into PLS, for example, eliminating the use of technical jargon, replacing passive voice by active, or using short sentences and paragraphs [6, 11]. However, authoring a PLS can be time consuming and challenging, particularly in areas like clinical settings which typically involve documents with technical and domain-specific vocabulary.

With the advancement of technology, new methods have been developed to automate the simplification of biomedical texts. In 2022, a review by Oldov et al. analyzed 32 tools or methods using either rule-based approach or Natural Language Processing (NLP) and concluded that NLP methods offer more promising outputs but were limited by scarcity of training data, resulting in continued reliance on rule-based methods [12]. Large Language Models (LLMs) with their immense data training potential and text generation capabilities, present a promising solution to tackle this challenge and automate the generation of PLS from technical documents.

With the objective of bridging the gap in health literacy by facilitating the translation of biomedical texts to comprehensible summaries designed for patients, our study demonstrates the potential of NLP to develop a classification system to identify if a text is written in plain language, and LLMs to automate the generation of accurate, complete, and comprehensible PLS.

## Materials and Methods

Our methodology, outlined in Figure 1, consisted of 3 main steps: collecting and processing of sample texts in technical and plain language, conducting a quantitative analysis of the texts to generate a plain language classification model and a qualitative analysis to generate the prompts for the LLMs, and using the LLMs to generate PLS and test them.

**Figure 1.**
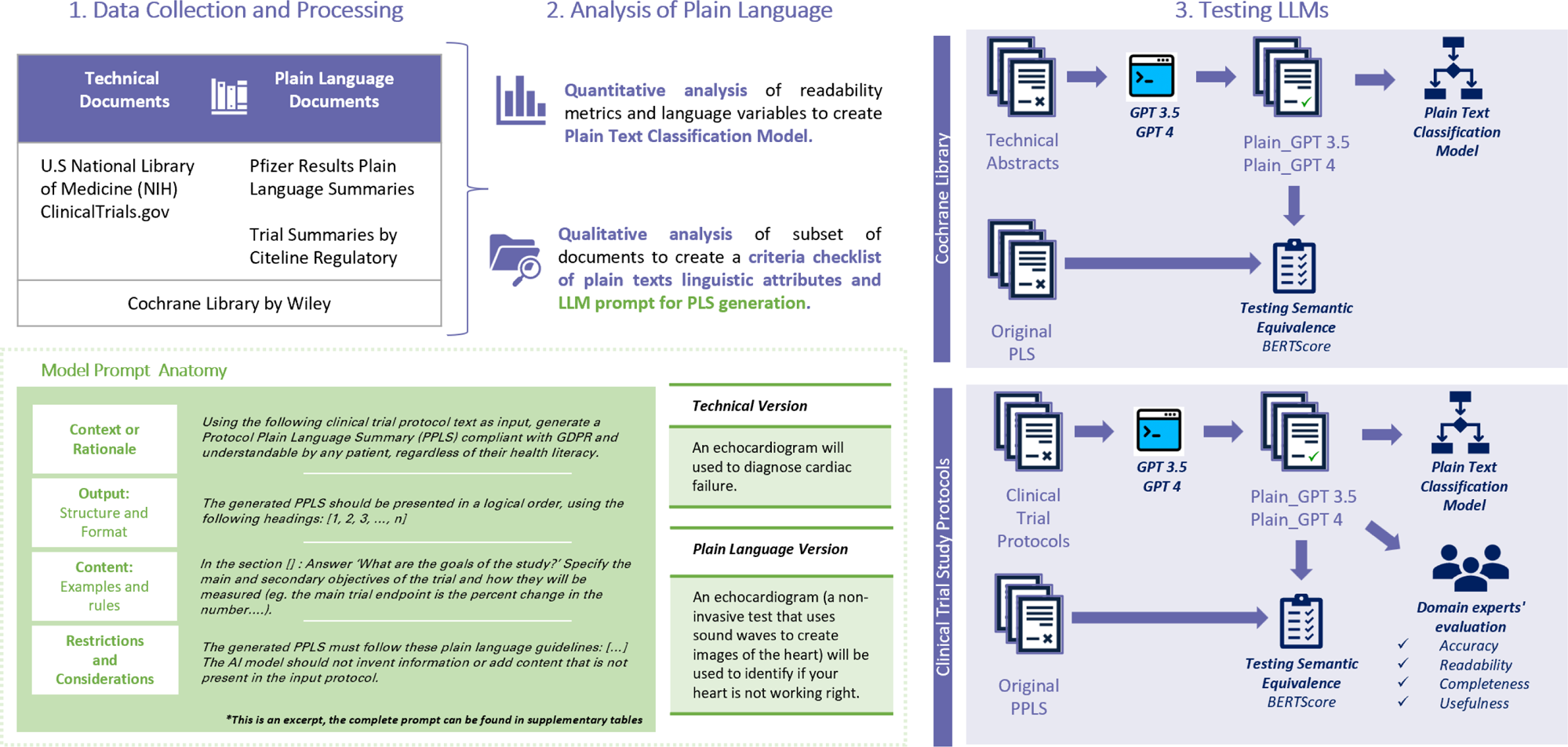
Methodology. Our methodology involved three steps: 1) collection and processing of biomedical texts (technical documents and plain language documents) into datasets for training and testing, 2) quantitative analysis of the texts to create a plain language classification model, and qualitative analysis to identify linguistic traits in plain texts to guide the engineering of a prompt that could translate biomedical text into Plain Language Summaries (PLS) using Language Learning Models (LLMs; and 3) testing the effectiveness of the LLMs in generating PLS quantitatively with our classification model and with semantic equivalence (BERTScore) and qualitatively with domain experts’ evaluation.

### Data Collection and Processing

We collected biomedical texts, both in technical and plain language (see the data sources in *Supplementary Table 1*) and assembled them into a dataset of 14,441 texts. This “*main dataset”* was then divided into training and testing sets, consisting of 4,596 plain and 6,721 technical texts for training, and 1,149 plain and 1,975 technical texts for testing.

We enlarged each dataset by treating each paragraph of a minimum of 250 words as a distinct unit, while excluding texts with fewer than 250 words. As a result, our “*augmented dataset*” had 61,354 texts, divided into 16,731 plain and 31,740 technical for training, and 5,090 plain and 7,793 technical for testing.

### Analysis of Plain Language

We conducted qualitative and quantitative analysis of the texts to identify unique linguistic traits and variables that classify a text as plain language.

#### Qualitative Analysis

Driven by the varying and broad-scope guidance on creating high-quality PLS [13], we analyzed a subset of our plain texts and created a ‘criteria checklist’ (*see Supplementary Table 2*) with the linguistic attributes most commonly present in plain texts. Key resources used in this process were guides and reviews, such as: Your Guide to CLEAR WRITING by CDC [11], Federal Plain Language Guidelines [14], Health Literacy Universal Precautions Toolkit by Agency for Healthcare Research and Quality (AHRQ) [15], Just Plain Clear Glossary by United Health Group [16], EU 536/2014 Summary of Clinical Results for Laypersons [17], and results presented by Stoll et al, in their systematic review of theory, guidelines, and empirical research on PLS [13]. We used the resultant checklist to complement the qualitative findings described in the next section and aid in developing the prompt detailed in the section LLM Prompt for Plain Language Summary Generation.

#### Quantitative Analysis

We computed readability metrics and language variables for each text in the augmented dataset using the Readability library [18] and SpaCy [19], respectively. This resulted in 64 variables presenting each text’s readability and linguistic traits (*see Supplementary Table 3*).

We analyzed the language variables in our dataset to identify their potential to classify a text as technical or plain. We used statistical hypothesis test for each of the variables of the *main dataset*. For each variable, we created a random sample of size *n* from the plain texts (*X*_1_, *X*_2_…*X*_*n*_ ∼ *P*_*X*_) and a random sample of size *n* from the technical texts (*Y*_1_, *Y*_2_…*Y*_*n*_ ∼ *Q*_*Y*_), and tested if our data supported either of the following hypotheses:

- *Null Hypothesis*, *H*_0_:*P* = *Q*, the distributions of the proportion of the variable of interest for both samples (text and technical) are the same.
- *Alternate Hypothesis*, *H*_0_:*P* ≠ *Q*, the distributions of the proportion of the variable of interest for both samples (text and technical) are different.

We evaluated the null hypothesis by comparing our 2 distributions using non-parametric tests: Wilcoxon, Kolmogorov-Smirnov (KS), and Mann–Whitney U. Given the multiple hypothesis tests, one for each variable, we adjusted the significance levels to control the probability of Type I errors by using the Bonferroni correction to lower the alpha value by dividing the desired significance level ∝= 0·05 by the total number of tests *m* = 64^18^.

Figure 2 illustrate examples of the comparison of the distributions of some of the variables in technical and plain texts. Out of the 64 variables, only ‘Interjections’ and ‘Passive Voice’ did not provide sufficient evidence to reject the null hypothesis (*ρ*-value > 0·0008). The other 62 variables were significantly distinct between the types of text and were included in our classification model.

**Figure 2.**
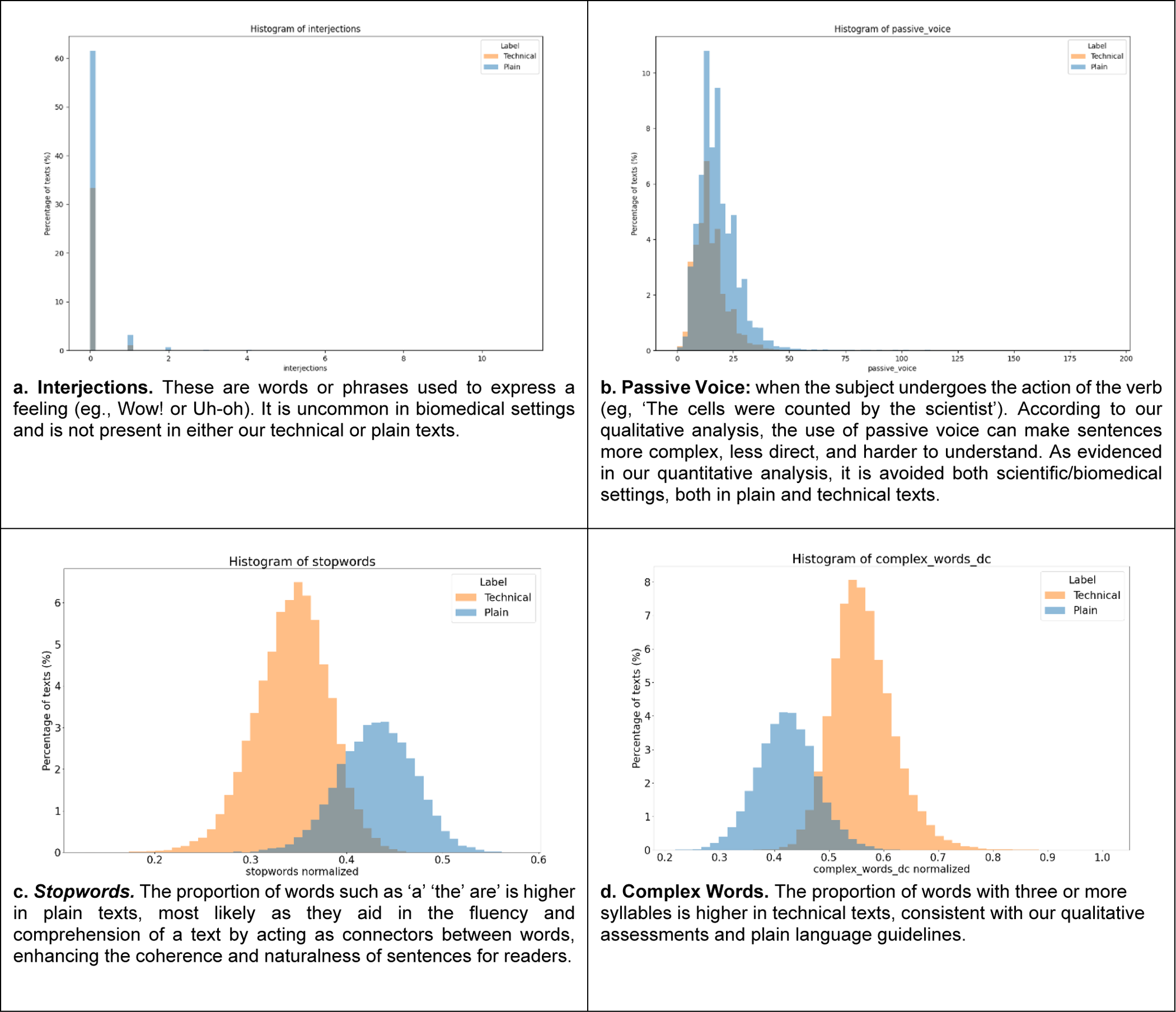
Comparison of the distribution of a sample of readability metrics or language variables between plain and technical texts.

### Plain Texts Classification Model

We used the *augmented dataset - train* and the 62 distinct variables between text types (Section Quantitative Analysis), to build the classification model. We used Gradient Boosting (GB) and Random Forest (RF) machine learning models.

### LLM Prompt for Plain Language Summary Generation

Our objective was to design a prompt for LLMs capable of translating biomedical technical documents into PLS.

Beginning with a clinical trial protocol from ClinicalTrials.Gov (see data sources in *Supplementary Table 1*), we used a simple initial prompt: ‘*Using the following clinical trial protocol text as input, create a plain language summary’.* We tested this prompt using both GPT3.5 and GPT4, analyzed the generated output, and iteratively refined the prompt by adding details and instructions.

We aimed to produce a PLS that met the following qualitative criteria: (1) **Accuracy:** the content is clinically and scientifically accurate. (2) **Readability**: the content is comprehensible by a lay person, as defined by the plain language criteria checklist (*Supplementary* **Error! Reference source not found.**). (3) **Completeness:** the content adheres with the expectations of a Protocol Plain Language Summary (PPLS) as specified by EU CTR No 536/2014 [16]. **(4) Usefulness:** the generated PLS can be used as a first version to draft the study PPLS.

Our final prompt, provided in *Supplementary Table 4*, was designed specifically to generate a PLS of a clinical trial protocol. It includes the following elements:

- **Context:** a clear rationale on why a PLS is needed for the given clinical trial protocol.
- **Output:** the desired structure and format for the generated summary, including the specific sections of the output.
- **Content:** the expected content within each section, with examples and rules to guide the generation process.
- **Restrictions:** limitations of the output (e.g., word count limitations, the inclusion of only the information provided in the original protocol, and adherence to the criteria checklist for plain language as set out in *Supplementary* **Error! Reference source not found.**).

After finalizing the prompt for generating a PPLS, we used the same approach to create a prompt to generate Cochrane Reviews PLS (see the description of this data source in *Supplementary Table 1*, and the prompt in *Supplementary Table 5)*.

We used our prompts with GPT 3.5 and GPT 4 to translate technical biomedical texts, Cochrane Reviews and Study Protocols, into their respective PLS: Cochrane PLS and Protocol PLS. We quantitatively tested the generated PLS for plainness and semantic equivalence. For PPLS, we also performed a qualitative assessment of the outputs by three experts in Clinical Trial Operations and Regulatory Medical Writing, who rated each GPT 3.5 and GPT 4 text on a 5-point Likert Scale (1-Strongly Disagree to 5-Strongly Agree). They evaluated the texts for accuracy, readability, completeness, and usefulness as defined in the section: *LLM Prompt for Plain Language Summary Generation*.

## Results

### Plain Text Classification Model

The classification models accurately distinguished whether an input text was plain or technical. The Gradient Boosting model showed slightly superior results with a precision rate of 97·2% (See Table 1).

**Table 1.** Comparison of tested classification models in terms of F1 Score or predictive performance, Accuracy, Recall, and Precision.

| Model | F1 Score<br>(Predictive Performance) | Accuracy | Recall | Precision |
| --- | --- | --- | --- | --- |
| Random Forest | 0.971 | 0.980 | 0.973 | 0.969 |
| Gradient Boosting | 0.975 | 0.982 | 0.977 | 0.972 |

### LLM Prompt for Plain Language Summary Generation

#### Cochrane Reviews: Plain Language Summaries

We randomly selected a sample of 600 Cochrane texts from the *main dataset*: 300 technical abstracts and the corresponding 300 plain summaries. We then used our prompt in both GPT 3.5 and GPT 4 to generate the plain language summary from the technical abstracts resulting in 300 Plain-GPT 3.5 and 300 Plain-GPT 4 summaries.

We tested the LLM-generated texts with our best model, Gradient Boosting, for plain language classification, and BERTScore to test semantic equivalence against the original Cochrane plain summaries.

Our model classified 96% of GPT 3.5 texts and 99·6% of GPT 4 texts as plain. Hence, our prompt is effective in generating PLSs that meet quantitative plain language requirements as defined in our classification model, with GPT 4 showing higher adherence.

The semantic equivalence score, BERTScore, confirmed both GPT 3.5 and GPT 4 successfully retained the original message. However, GPT 4 produced plain summaries that outperformed GPT 3.5 in all parameters (Precision, Recall, and F1-Score) with a significant difference (*ρ*-value < 0.05) (Table 2).

**Table 2.** Semantic equivalence score (BERT) between the GPT-generated plain summaries from Cochrane technical abstract vs. original Cochrane PLS.

| Semantic<br>Equivalence | *Plain_GPT 3.5* | *Plain_GPT4* | *p-value* |
| --- | --- | --- | --- |
| Precision | 0.790 ± 0.010 | 0.791 ± 0.015 | 0.027 |
| Recall | 0.772 ± 0.017 | 0.773 ± 0.016 | 0.003 |
| F1-Score | 0.780 ± 0.015 | 0.782 ± 0.014 | 0.001 |

#### Protocol Plain Language Summaries

We randomly selected a sample of nine clinical trial protocols from ClinicalTrials.Gov. Given that their corresponding PPLS were not yet publicly published, we used Trial Summaries by Citeline Regulatory to find the corresponding Results Plain Language Summaries (RPLS) and extracted four sections that are equivalent in a PPLS: ‘Why is this study needed?’: Background and hypothesis of the trial (*Rationale*), ‘Who will take part in this study?’ (*Population*), ‘How is this study designed?’ (*Trial Design*), and ‘What treatments are being given during the study?’ (*Interventions*).

##### Quantitative Analysis

We used our prompt specific for PPLS with both GPT 3.5 and GPT 4 to generate the plain language summary from the technical protocols. We used our Gradient Boosting model to verify if LLM-generated texts were plain and BERTScore to check semantic equivalence to the content on the RPLS. All LLM-generated PPLS were classified as plain, and BERTScore confirmed semantic agreement with the content in the RPLS (Table 3). Consistent with Cochrane results, GPT 4 produced PPLS with higher semantic equivalence than GPT 3.5 (no statistical analysis due to small sample size).

**Table 3.**
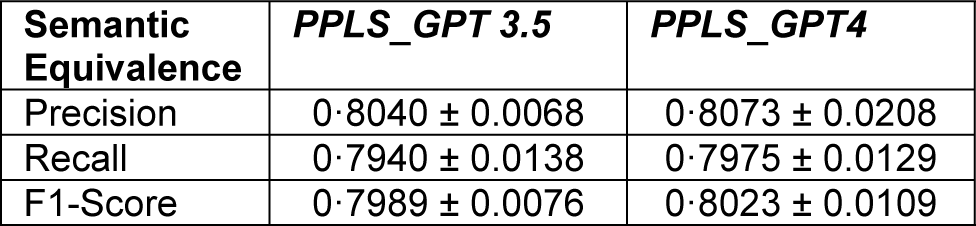
Semantic equivalence score (BERT) between the GPT-generated PPLS from clinical trial protocols vs. the original content written for the PLS.

##### Qualitative Analysis

Ratings by 3 domain experts who evaluated each LLM-generated text, demonstrated that GPT 4 outperformed GPT 3.5 in all four criteria: accuracy, readability, completeness and usefulness, as indicated by an average score of 4·71 for GPT 4 texts as compared to 3·93 for GPT 3.5 (see Figure 3 and Table 4).

**Figure 3.**
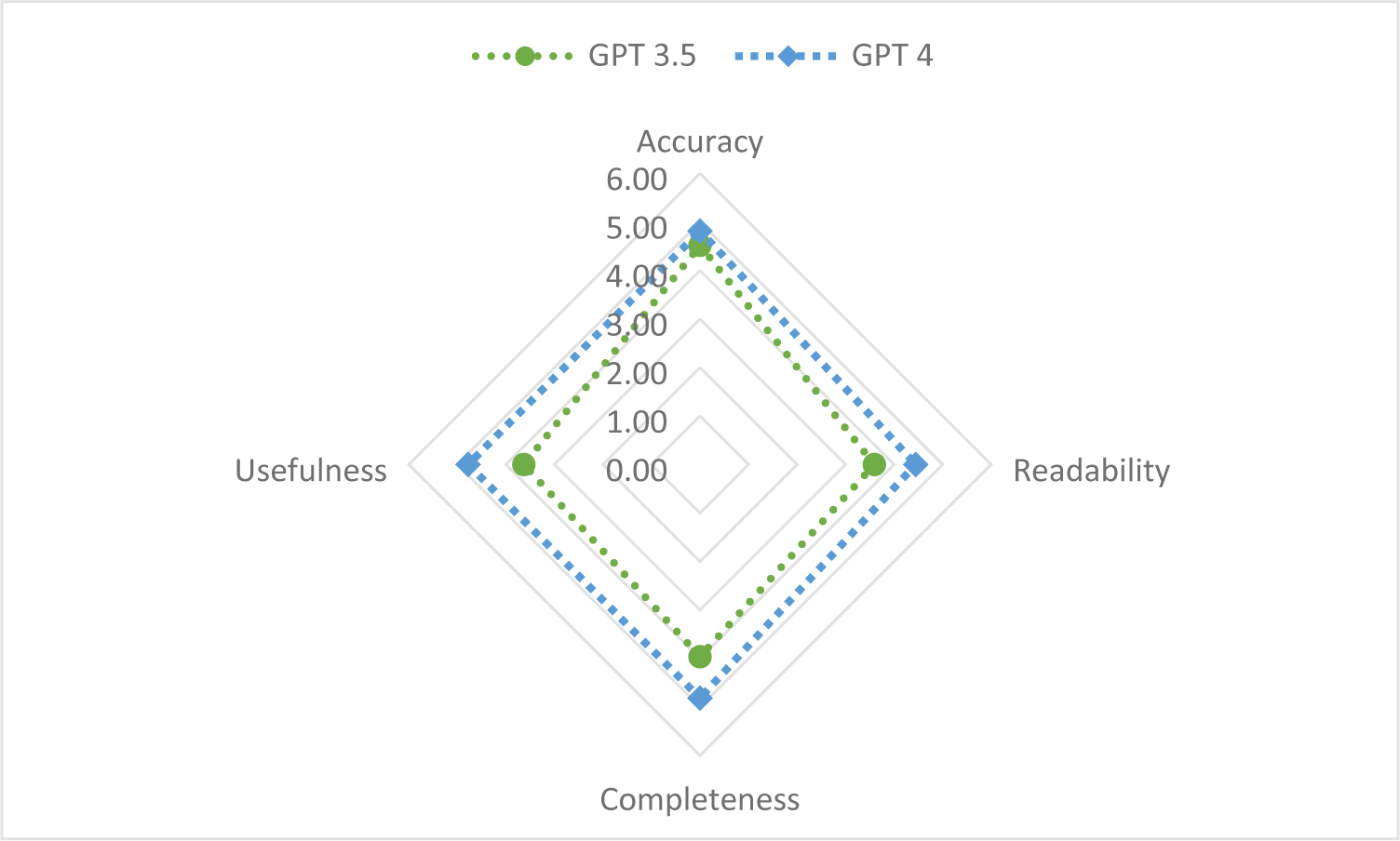
Radar diagram comparing the qualitative assessment of the LLM-generated texts in 4 criteria: Accuracy, Readability, Completeness, and Usefulness.

**Table 4.** Ratings for GPT 3.5 and GPT 4 plain summaries in 4 criteria: Accuracy, Readability, Completeness, and Usefulness.

|  | Accuracy | Readability | Completeness | Usefulness | Overall Score |
| --- | --- | --- | --- | --- | --- |
| **GPT 3.5** | 4.52 | 3.59 | 3.96 | 3.63 | **3.93** |
| **GPT 4** | 4.81 | 4.44 | 4.81 | 4.78 | **4.71** |

In terms of accuracy, both GPT 3.5 and GPT 4 received high scores. Reviewers noted that both language models exhibited scientific accuracy and relied exclusively on the input text (study protocol). Notably, even when the content in the original RPLS contained inconsistencies (e.g. incorrect age limit or indication), both language models generated accurate PLS. This finding suggests that language models can be used to automatically generate a first draft of a PLS while minimizing data inaccuracies resulting from human error.

Regarding readability, both GPT 3.5 and GPT 4 generated texts that were likely to be understood by a lay audience. This observation aligned with the results obtained through the classification model. However, GPT 3.5 occasionally employed complicated medical jargon (e.g., ‘chronic’, ‘randomized’, ‘double-blind’) and longer words and sentences (e.g., ‘approximately 640 adults’ vs ‘about 640 adults’). Similarly, GPT 4, despite its outstanding performance, occasionally prefered passive voice over active voice, compromising clarity and consice writing. This highlights the importance of quality control by a healthcare professional who should verify the content and style of the automatically generated PLS draft.

Completeness, which assessed the compliance of PPLS content and structure with EU CTR No 536/2014 guidelines, revealed inconsistencies in the outputs generated by GPT 3.5. These inconsistencies manifested as the creation of new, unrequested sections and summaries, with significant variation among the nine generated PLS. Conversely, GPT 4 consistently generated PLSs that adhered to the specified format and content expectations, and complied with the guidelines, showing a remarkable value in automating the time-consuming task of guaranteeing the content to be standardized and aligned with industry specific and rigorous guidelines.

The usefulness ratings, indicating the suitability of the generated PLSs as draft versions, correlated with the findings in other criteria. GPT 3.5 received moderate scores in generating draft PLS, while GPT 4 scored 4·78, indicating that the generated PLS were highly suitable as draft versions of the PLS.

## Discussion

In this study, we used NLP and LLMs to improve health literacy by generating PLS from biomedical texts. Our two-part strategy involved creating a classification model for identifying if a text was written in plain language, and using LLMs (specifically GPT 3.5 and GPT 4) for the automated generation of the PLS.

The classification model achieved over 97% accuracy, indicating its effectiveness in distinguishing between the text types: technical and plain. This is a very useful stand-alone strategy which could support authoring teams in identifying if their texts targeted for patients or the general audience are compliant with plain language guidelines.

The LLMs exhibited outstanding performance in generating PLS, with GPT 4 outperforming GPT 3.5 in creating content that was both plain and semantically similar. In a qualitative review by domain experts, GPT 4 also surpassed GPT 3.5 by generating high-quality drafts of PLS. These drafts were scientifically accurate, compliant with plain language requirements, and met expectations in content and structure. These results underlines the value of LLMs in supporting healthcare stakeholders to streamline the generation of plain documents, and with that, promote equitable access to biomedical information, engagement of the lay audience in health-related decision making, and improved health outcomes.

Our study highlights the importance of using well-designed, structured, and domain-specific prompts to guarantee the creation of high-quality, easily comprehensible PLS. This is particularly vital when accuracy in biomedical facts is essential. This requires the collection of feedback from stakeholders who are experts in the domain or field of interest. Such feedback would help to fine-tune the prompts and guarantee that the output fullfils the purposes of different document types. Our study exemplified this with various document types (e.g., Cochrane reviews, PPLS), some of which adhere to strict industry standards.

While the findings of our study are promising, they also underscore opportunities for further research to fully harness the potential of NLP and LLMs in this context. Future studies could involve direct audience feedback in evaluating the understandability of PLS. This would ensure that the generated content aligns with the comprehension levels of the intended audience, such as patients in clinical settings, and would provide cues for ways in which the they could improve their interaction with biomedical content, improving adherence to treatment plans or educating them about a disease or diagnosis. Additionally, depending on the intended use and field of interest, refining the models could potentially account for specific linguistic nuances, exploring advanced techniques like Retrieval Augmented Generation (RAG) could enhance factual accuracy, and expanding the dataset to include a wider range of texts and languages could enhance the generalizability of the classification model and applicability of the LLMs. Different interesting opportunities to leverage NLP and LLMs to serve society by simplifying what would otherwise be daunting.

In conclusion, by leveraging the capabilities of NLP and LLMs, we have taken a significant step towards bridging the gap between complicated biomedical texts and comprehensible summaries designed for the general audience. This framework paves the way for prospective innovations in the field of health literacy, which, in turn, holds the potential to enhance health outcomes and foster health equity.

## Data Availability

Data is fully available, without restriction. All data sources are listed in the supplemental appendix and include a link to the GitHub Data Repository.

https://github.com/feliperussi/bridging-the-gap-in-health-literacy/tree/main

